# Direct detection of SARS-CoV-2 using CRISPR-Cas13a and a mobile phone

**DOI:** 10.1101/2020.09.28.20201947

**Authors:** Parinaz Fozouni, Sungmin Son, María Díaz de León Derby, Gavin J. Knott, Carley N. Gray, Michael V. D’Ambrosio, Chunyu Zhao, Neil A. Switz, G. Renuka Kumar, Stephanie I. Stephens, Daniela Boehm, Chia-Lin Tsou, Jeffrey Shu, Abdul Bhuiya, Max Armstrong, Andrew Harris, Jeannette M. Osterloh, Anke Meyer-Franke, Charles Langelier, Katherine S. Pollard, Emily D. Crawford, Andreas S. Puschnik, Maira Phelps, Amy Kistler, Joseph L. DeRisi, Jennifer A. Doudna, Daniel A. Fletcher, Melanie Ott

## Abstract

The December 2019 outbreak of a novel respiratory virus, SARS-CoV-2, has become an ongoing global pandemic due in part to the challenge of identifying symptomatic, asymptomatic and pre-symptomatic carriers of the virus. CRISPR-based diagnostics that utilize RNA and DNA-targeting enzymes can augment gold-standard PCR-based testing if they can be made rapid, portable and accurate. Here we report the development of an amplification-free CRISPR-Cas13a-based mobile phone assay for direct detection of SARS-CoV-2 from nasal swab RNA extracts. The assay achieved ∼100 copies/μL sensitivity in under 30 minutes and accurately detected a set of positive clinical samples in under 5 minutes. We combined crRNAs targeting SARS-CoV-2 RNA to improve sensitivity and specificity, and we directly quantified viral load using enzyme kinetics. Combined with mobile phone-based quantification, this assay can provide rapid, low-cost, point-of-care screening to aid in the control of SARS-CoV-2.

## INTRODUCTION

In late 2019, a novel infectious respiratory RNA virus, severe acute respiratory syndrome (SARS)-coronavirus (CoV)-2, emerged in the human population, likely from a zoonotic source (Wang et al., 2020; Zhu et al., 2020). In most people, SARS-CoV-2 infection causes mild or no symptoms. Critically, however, asymptomatic or lowly symptomatic carriers spread the virus (Lee et al., 2020), leading to delayed isolation of carriers and massive worldwide spread (Bai et al., 2020; Lavezzo et al., 2020).

The current gold-standard diagnostic for SARS-CoV-2 infection, quantitative reverse transcription polymerase chain reaction (RT-qPCR), is well established and widely used for screening. Based on primers directed against the nucleocapsid (N), envelope (E), and open reading frame 1ab (ORF1ab) genes, RT-qPCR has an analytical limit of detection (LOD) of 1,000 viral RNA copies/mL (1 copy/μL) (Vogels et al., 2020). However, recent modeling of viral dynamics suggests that frequent testing with a fast turn-around time is required to break the current pandemic (Larremore et al., 2020). Notably, the model ranked sensitivity of the test as a lower priority, and estimated that an LOD of 100,000 copies/mL (100 copies/μL) would be sufficient for screening (Larremore et al., 2020). In clinical studies, when the viral load drops below a million copies/mL (1,000 copies/μL), few infectious particles are detected and consequently the risk of viral transmission is low (La Scola et al., 2020; Quicke et al., 2020; Wolfel et al., 2020). Since the high sensitivity of RT-qPCR may pick up RNAs freed by infected and dead cells after infectious particles have waned, use of qPCR for screening may lead to unnecessary isolation of individuals that are SARS-CoV-2 RNA-positive but no longer infectious (Alexandersen et al., 2020).

The need for SARS-CoV-2 tests that are rapid, widespread, and able to identify infectious individuals has motivated efforts to explore new strategies for viral RNA detection based on CRISPR technology. Cas12 and Cas13 proteins are RNA-guided components of bacterial adaptive immune systems that directly target single- and double-stranded DNA or single-stranded (ss)RNA substrates, respectively (Abudayyeh et al., 2016; Chen et al., 2018; East-Seletsky et al., 2016; Zetsche et al., 2015). Cas13 is complexed with a CRISPR RNA (crRNA) containing a programmable spacer sequence to form a nuclease-inactive ribonucleoprotein complex (RNP). When the RNP binds to a complementary target RNA, it activates the HEPN (higher eukaryotes and prokaryotes nucleotide-binding domain) motifs of Cas13 that then indiscriminately cleave any surrounding ssRNAs. Target RNA binding and subsequent Cas13 cleavage activity can therefore be detected with a fluorophore-quencher pair linked by an ssRNA, which will fluoresce after cleavage by active Cas13 (East-Seletsky et al., 2016). To date, four type VI CRISPR-Cas13 subtypes have been identified: Cas13a (previously known as C2c2) (Abudayyeh et al., 2016; East-Seletsky et al., 2016; Shmakov et al., 2015), Cas13b (Smargon et al., 2017), Cas13c (Shmakov et al., 2017), and Cas13d (Konermann et al., 2018; Yan et al., 2018).

What initially evolved as a successful strategy in bacteria to destroy incoming phages and build a immunological memory is now being harnessed for viral diagnostics (Chen et al., 2018; East-Seletsky et al., 2016; Gootenberg et al., 2018; Gootenberg et al., 2017; Myhrvold et al., 2018). To achieve high sensitivity, current CRISPR diagnostic (CRISPR Dx) strategies rely on pre-amplification of target RNA for subsequent detection by a Cas protein. In the case of the RNA-sensing Cas13 proteins, this typically entails the conversion of RNA to DNA by reverse transcription, DNA-based amplification (i.e., isothermal amplification, loop-mediated isothermal amplification), and transcription back to RNA for detection by Cas13a or Cas13b, called “SHERLOCK” (Gootenberg et al., 2018; Gootenberg et al., 2017). This was recently adapted for SARS-CoV-2 RNA detection (Joung et al., 2020), and further developed as “SHINE” for testing unextracted samples (Arizti-Sanz et al., 2020). The conversion of amplified DNA back into RNA can be avoided by using the DNA-sensing Cas12 for detection, a method called “DETECTR” (Chen et al., 2018), which has recently been adapted for SARS-CoV-2 detection (Broughton et al., 2020). Both SHERLOCK and DETECTR reactions take approximately an hour to complete and can be read out with paper-based lateral flow strips appropriate for point-of-care use, although their current FDA-approved protocols are still laboratory-based.

Here, we report the development and demonstration of a rapid CRISPR-Cas13a-based assay for the direct detection of SARS-CoV-2 RNA. This assay, unlike previous CRISPR diagnostics, does not require pre-amplification of the viral genome for detection. By directly detecting the viral RNA without additional manipulations, the test yields quantitative RNA measurements rather than simply a positive or negative result. To demonstrate the simplicity and portability of this assay, we measure fluorescence with a mobile phone camera in a compact device that includes low-cost laser illumination and collection optics. The high sensitivity of mobile phone cameras, together with their connectivity, GPS and data-processing capabilities, have made them attractive tools for point-of-care disease diagnosis in low-resource regions (Breslauer et al., 2009; D’Ambrosio et al., 2015; Kamgno et al., 2017; Wood et al., 2019). By combining multiple crRNAs to increase Cas13a activation and analyzing the change in fluorescence over time rather than solely endpoint fluorescence, we are able to achieve SARS-CoV-2 viral RNA detection of ∼100 copies/μL within 30 minutes. We also correctly identified all SARS-CoV-2 positive patient samples tested (Ct values 14.37 to 22.13) within 5 minutes. This approach has the potential to enable a fast, accurate, portable, and low-cost option for point-of care SARS-CoV-2 screening.

## RESULTS

### Quantitative Direct Detection of Viral SARS-CoV-2 RNA with Cas13a

When the SARS-CoV-2 sequence became public in January 2020, we set out to develop a Cas13-based direct-detection assay for viral RNA that would avoid the need for amplification and enable point-of-care testing. To do this, we needed to optimize Cas13 activation through careful crRNA selection and develop a sensitive and portable fluorescence detection system for our assay (Figure 1A). Initially, we designed 12 crRNAs along the nucleocapsid (N) gene of SARS-CoV-2, corresponding to the three Centers for Disease Control and Prevention (CDC) N primer sets and the N primer set developed early in the pandemic in Wuhan, China (Zhu et al., 2020). Every Cas13-crRNA RNP should detect a single 20-nucleotide region in the N gene (Figure 1B).

**Figure 1:**
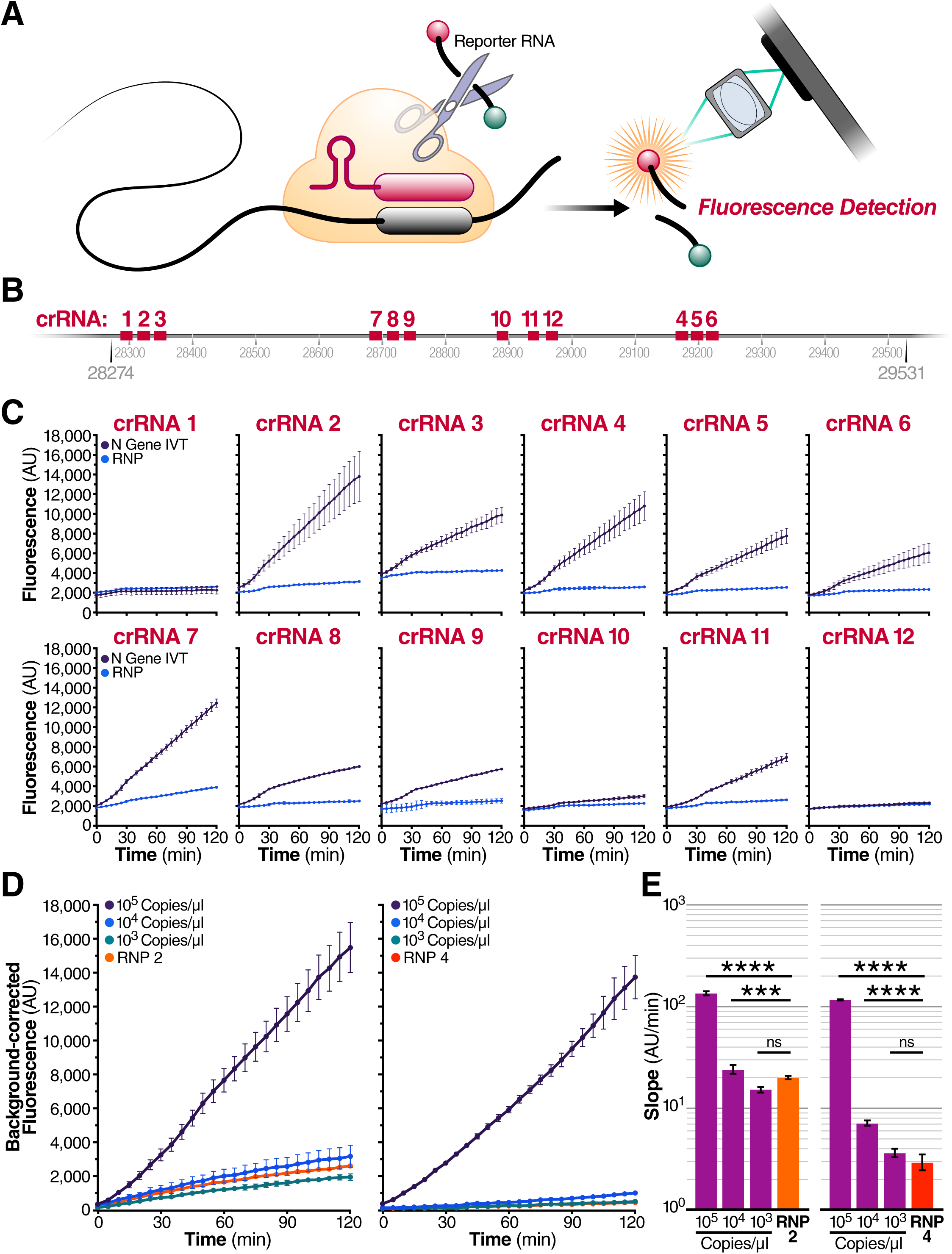
Quantitative Direct Detection of Viral SARS-CoV-2 RNA with Cas13a. (A) Schematic of a Cas13a (beige)-crRNA (red) RNP complex binding target RNA (black), resulting in activation of the HEPN nuclease (denoted by scissors) domain. Upon target recognition and RNP activation, Cas13a indiscriminately cleaves a quenched-fluorophore RNA reporter, allowing for fluorescence detection as a proxy for Cas13a activation and target RNA. (B) Schematic of the SARS-CoV-2 nucleocapsid (N) gene, and the corresponding locations of each crRNA spacer region. (C) Cas13a RNPs made individually with each crRNA (final RNP complex concentration of 50 nM) were tested against 2.9 x 10^5^ copies/μL (480 fM) of SARS-CoV-2 *in vitro* transcribed N gene RNA in a total 20 μL reaction volume. Background fluorescence by the individual RNP in the absence of target RNA is shown as “RNP.” Raw fluorescence values over two hours is shown. Data are represented as mean ± standard deviation (SD) of three technical replicates. (D) Limit of detection of crRNA 2 and crRNA 4 was determined by testing each 100 nM of each RNP individually against 10^5^, 10^4^, and 10^3^ copies/μL of N gene IVT RNA. “RNP 2” and “RNP 4” represent no target RNA RNP alone controls. Background correction of fluorescence was performed by subtraction of reporter alone fluorescence values. Data are represented as mean ± standard error of the difference between means of three technical replicates. (E) Slope of the curve over two hours from Figure 1D was calculated by simple linear regression and shown as slope ± 95% confidence interval. Slopes were compared to the RNP alone control through an Analysis of Covariance (ANCOVA): ∗∗∗∗p<0.0001, ∗∗∗p<0.001, ns=not significantly higher than RNP control.

We first tested each crRNA individually in a direct-detection assay on a plate reader. We selected the Cas13a homolog from *Leptotrichia buccalis* (Lbu) as it demonstrated the highest sensitivity and robust *trans*-cleavage activity relative to other characterized Cas13a homologs (East-Seletsky et al., 2017; East-Seletsky et al., 2016). The assay used purified LbuCas13a (East-Seletsky et al., 2017; East-Seletsky et al., 2016) and a quenched fluorescent RNA reporter (East-Seletsky et al., 2017; East-Seletsky et al., 2016), together with *in vitro* transcribed (IVT) target RNA corresponding to the viral N gene (nucleotide positions 28274–29531). At a target RNA concentration of 480 fM (2.89 x 10^5^ copies/μl), we identified 10 crRNAs with reactivity above the RNP alone control that contained the same RNP and probe but no target RNA (Figure 1C). The use of RNase-free buffers minimized background fluorescence, and the plate reader gain and filter bandwidth settings were optimized to capture low-level reporter cleavage. Similar results were obtained when full-length viral RNA was used as target RNA (Supplemental Figure 1A). For the initial studies, we selected two crRNAs (crRNAs 2 and 4) that generated the greatest Cas13a activation as determined by the fluorescent reporter while maintaining low levels of target-independent fluorescence (indicated by the RNP alone curve).

We next carried out serial dilutions of the target RNA to independently determine the limit of detection for each crRNA. LbuCas13a exhibited detectable reporter cleavage with as little as 10 fM (∼6000 copies/μL) of target RNA (East-Seletsky et al., 2017). Consistent with this, we found that RNPs made with either crRNA 2 and crRNA 4 did not appear to generate signals above the RNP controls for an IVT target RNA concentration of 1,000 copies/μL (Figure 1d). The signal generated by direct detection with Cas13a appeared proportional to the concentration of target RNA in the assay. Given that the signal generated depends solely on the RNase activity of Cas13a, the linear rate of the reaction should approximate Michaelis-Menten enzyme kinetics. To determine if our assay was indeed quantitative, we compared the slopes determined by linear regression for different target RNA concentrations, using 1 to 10 µM KM and 600/s Kcat for the modeling (Slaymaker et al., 2019) (Figure 1E). The results confirmed that crRNA 2 and 4 each facilitated detection of at least 10,000 copies/μL of IVT N gene RNA. Since the measured slopes are proportional to the concentration of activated Cas13a, we could confirm that the rate of Cas13a activity scaled with concentration of target RNA (Supplementary Figure 2). This ability to estimate target RNA concentration from the measured slope allows for direct quantification of viral load in unknown samples.

### Combining crRNAs Improves Sensitivity of Cas13a

We next wondered whether combining crRNAs to form two different populations of RNPs in the same reaction could enhance overall Cas13a activation and, therefore, the sensitivity of the direct detection assay. In theory, a single target RNA could activate multiple Cas13a RNPs if each RNP is directed to different regions of the same viral target RNA, effectively doubling the active enzyme concentration (Figure 2A). To test this, we combined crRNAs 2 and 4 in the same reaction, keeping the total concentration of Cas13a RNPs constant but divided equally between RNPs made with each crRNA. We found that combining crRNA 2 and 4 markedly increased the slope of the detection reaction and the sensitivity of the reaction when measured with a fixed IVT target RNA concentration (480 fM) (Figure 2B). The slope increased from 213 AU/min (SE ± 1.6) (crRNA 2) and 159 AU/min (SE ± 1.7) (crRNA 4), individually, to 383 AU/min (SE ± 3.0) in combination, without increasing the slope of the RNP control reactions. This nearly doubling of the average slope compared to the individual crRNA reactions demonstrates the advantage of crRNA combinations.

**Figure 2:**
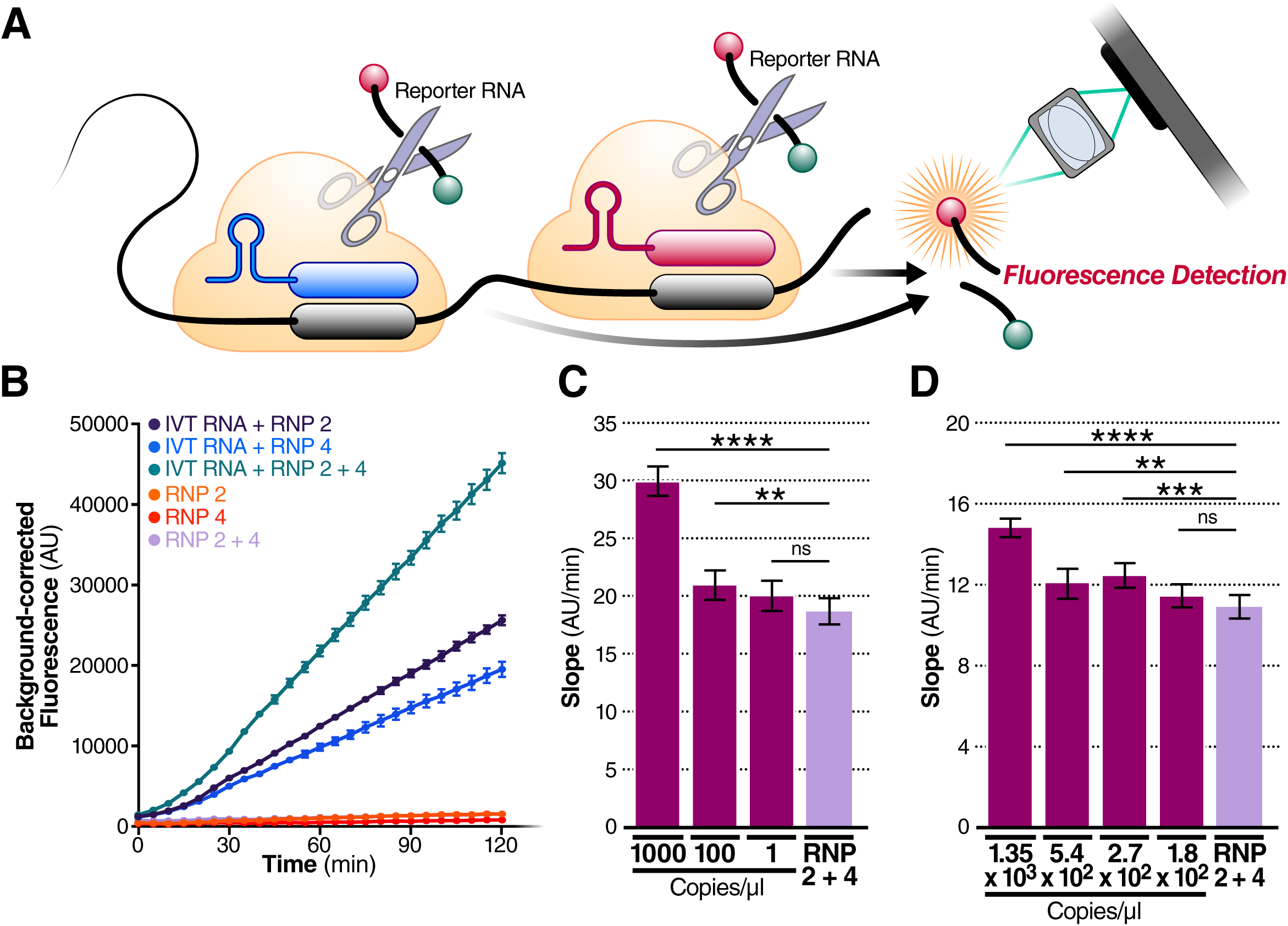
Combining crRNAs Improves Sensitivity of Cas13a. (A) Schematic of two different RNPs binding to different locations of the same SARS-CoV-2 RNA, leading to cleavage of the RNA reporter and increased fluorescence. (B) RNPs made with crRNA 2 and crRNA 4 individually and in combination (50 nM total RNP concentration for each reaction) were tested against 2.9 x 10^5^ copies/μL (480 fM) of SARS-CoV-2 IVT N gene RNA, and compared to fluorescence from no target RNA RNP alone controls (“RNP 2,” “RNP 4,” and “RNP 2+4”). Background correction of fluorescence was performed by subtraction of reporter alone fluorescence values. Data are represented as mean ± standard error of the difference between means of three technical replicates. (C) Limit of detection of crRNA 2 and crRNA 4 in combination was determined by combining 50 nM of RNP 2 and 50 nM of RNP 4 (100 nM total RNP) against 1,000, 100, and 1 copy/μL of SARS-CoV-2 IVT RNA (n=3, technical replicates). Slope of the curve over two hours was calculated by simple linear regression and is shown as slope ± 95% confidence interval. Slopes were compared to the no target RNA RNP alone control using ANCOVA: ∗∗∗∗p<0.0001, ∗∗p=0.0076, ns=not significant. (D) Limit of detection of crRNA 2 and crRNA 4 in combination was determined by combining 50 nM of RNP 2 and 50 nM of RNP 4 (100 nM total RNP) against 1.35 x 10^3^, 5.4 x 10^2^, 2.7 × 10^2^, and 1.8 × 10^2^ copies/μL of SARS-CoV-2 full-length viral RNA as quantified by qPCR (n=3, technical replicates). Slope of the curve over two hours was calculated by simple linear regression and is shown as slope ± 95% confidence interval. Slopes were compared to the no target RNA RNP alone control using ANCOVA: ∗∗∗∗p<0.0001, ∗∗∗p=0.0002, ∗∗p=0.0023, ns=not significant.

To determine how crRNA combinations affect the limit of detection, we tested crRNA 2+4 with a dilution series of IVT N gene RNA. The RNP combination improved the limit of detection, compared to the no-target RNP control (RNP 2+4), below 1,000 copies/μL of IVT target RNA (Figure 2C). We performed the same assay with full-length SARS-CoV-2 RNA isolated from the supernatant of SARS-CoV-2-infected Vero CCL-81 cells and found the limit of detection was 270 full-length viral copies/μL (Slope 12.4 ± SE 0.3) (Figure 2D). Viral copy numbers were determined by standard RT-qPCR. The difference between the viral IVT and full genome limits of detection could be explained by different quantification techniques of the target RNA or by considerable secondary structure in the viral RNA (Manfredonia et al., 2020; Sanders et al., 2020) that reduces RNP on-rates, thereby lowering the affinity of the RNP for the target (Abudayyeh et al., 2016).

One major advantage of CRISPR diagnostics is that they can be highly specific. To confirm the specificity of our crRNAs, we tested them against a set of other respiratory viruses, including alphacoronavirus HCoV-NL63 and betacoronaviruses HCoV-OC43 and Middle East respiratory syndrome coronavirus (MERS-CoV), which are among seven coronaviruses that infect human hosts and cause respiratory diseases (Fung and Liu, 2019). We extracted RNA from the supernatant of Huh 7.5.1-ACE2 or Vero E6 cells infected with HCoV-NL63 or HCoV-OC43, respectively, and produced IVT N gene RNA from MERS-CoV. In our Cas13a direct-detection assay with crRNA 2 and 4, individually and in combination, we detected no signal above RNP background for any of viral RNAs tested (Figure 3A). Similarly, no signal was detected with H1N1 Influenza A or Influenza B viral RNA, or with RNA extracted from primary human airway organoids (Figure 3B).

**Figure 3:**
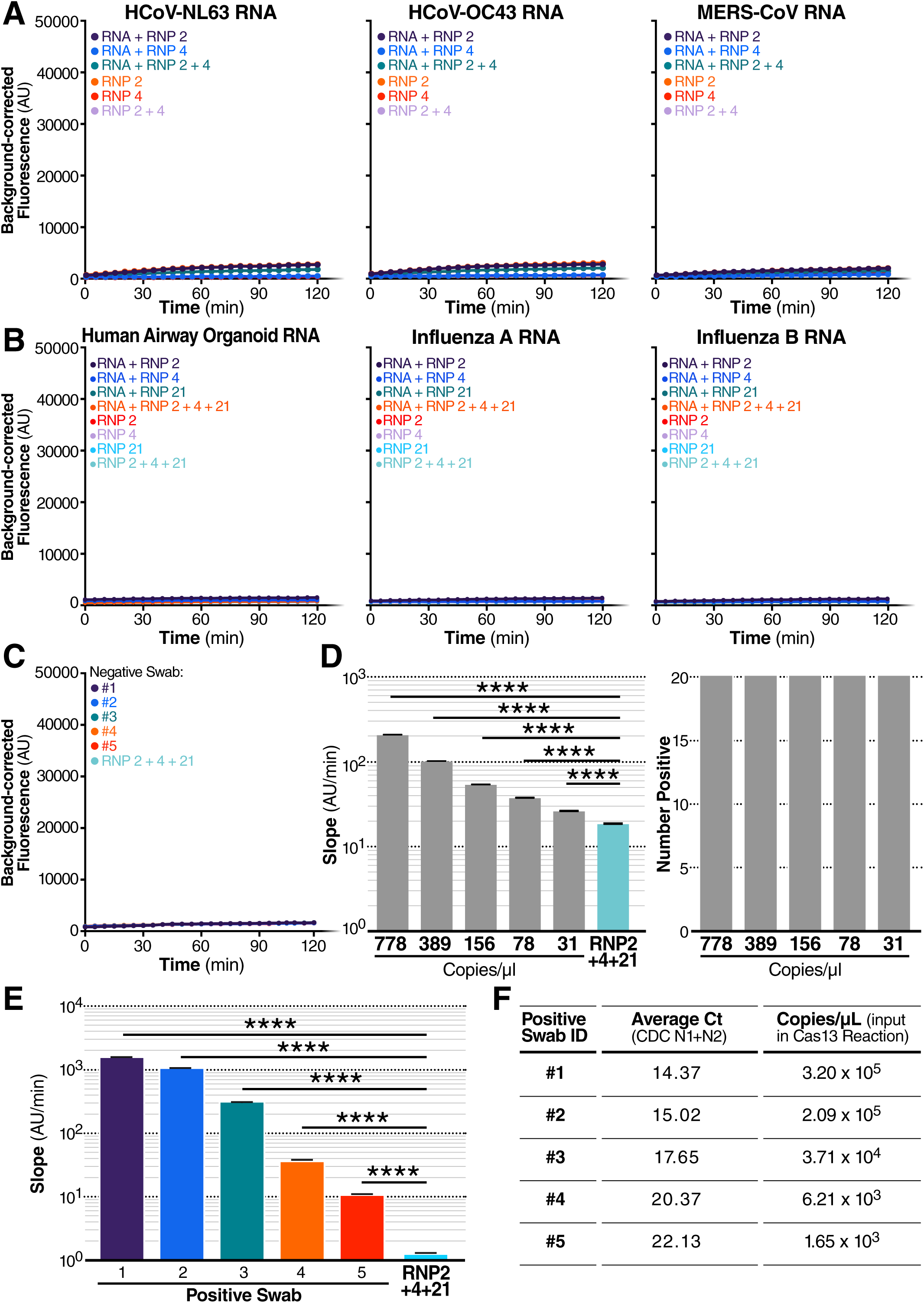
Cas13a Directly Detects SARS-CoV-2 RNA in Patient Samples. (A) crRNA 2 and crRNA 4 were tested individually (100 nM total RNP concentration) and in combination (100 nM total RNP concentration: 50 nM each of RNP 2 and RNP 4) against RNA isolated from HCoV-NL63 viral supernatant (left) and HCoV-OC43 viral supernatant (center) or the IVT N gene RNA from MERS-CoV (right). No target RNA RNP alone controls are denoted as “RNP 2,” “RNP 4,” and “RNP 2+4.” Background correction of fluorescence was performed by subtraction of reporter alone fluorescence values. Data are represented as mean ± standard error of the difference between means of three technical replicates. (B) crRNA 2 and crRNA 4 and crRNA 21 (see also Supplementary Figure 2) were tested individually (100 nM total RNP concentration) and in combination (100 nM total RNP concentration: 33 nM each of RNP 2, RNP 4, and RNP 21) against RNA isolated from human airway organoids (left), H1N1 Influenza A (center), and Influenza B (right). No target RNA RNP alone controls are denoted as “RNP 2,” “RNP 4,” “RNP 21,” “RNP 2+4+21.” Background correction of fluorescence was performed by subtraction of reporter alone fluorescence values. Data are represented as mean ± standard error of the difference between means of three technical replicates. (C) RNA from 5 nasopharyngeal swabs confirmed negative for SARS-CoV-2 by RT-qPCR was tested against RNP 2+4+21 (100 nM total RNP concentration). The no target RNA RNP control is denoted as “RNP 2+4+21.” Background correction of fluorescence was performed by subtraction of reporter alone fluorescence values. Data are represented as mean ± standard error of the difference between means of three technical replicates. (D) Dilutions of full-length SARS-CoV-2 RNA independently quantified by BEI using ddPCR was tested against RNP 2+4+21 to determine the limit of detection (n=20, technical replicates). Slope of the raw fluorescence curve over two hours was calculated by simple linear regression and is shown as slope ± SEM (left). Slopes were compared to the no target RNA RNP alone control using ANCOVA: ∗∗∗∗p<0.0001. An individual reaction containing the diluted SARS-Cov-2 RNA was compared with the reaction without the target RNA and the number of true positives was counted at the 95% confidence level (right). (E) RNA from 5 nasopharyngeal swabs confirmed positive for SARS-CoV-2 by RT-qPCR was tested against RNP 2+4+21 (100 nM total RNP concentration) (n=3, technical replicates). The no target RNA RNP alone control is denoted as “RNP 2+4+21.” Slope of the raw fluorescence curve over two hours was calculated by simple linear regression and is shown as slope ± 95% confidence interval. Slopes were compared to the no target RNA RNP background control using ANCOVA: ∗∗∗∗p<0.0001. (F) The Ct value (average Ct count using CDC N1 and N2 primers in RT-qPCR and the copies/μL input into the Cas13a reaction (See Figure 1E) are described for the RNA samples from each positive swab used in Figure 1E.

### Cas13a Directly Detects SARS-CoV-2 RNA in Patient Samples

We next examined whether our assay could be used with patient samples, where the swab and patient matrix (e.g., mucous from a nasal swab) could contribute additional background signal and reduce sensitivity. To increase assay performance prior to testing patient samples, we examined an additional set of crRNAs (crRNAs 19-22) targeting the viral E gene, based on published PCR primer and Cas12 guide sets (Corman et al., 2020) (Broughton et al., 2020) (Supplementary Figure 3A). When tested against full-length SARS-CoV-2 RNA, the crRNA that performed best, both individually (Supplementary Figure 3B) and in combination with crRNA 2 and 4, was crRNA 21 (Figure 3D). When tested on RNA from five SARS-CoV-2-confirmed negative nasal swab samples, the triple combination (RNP 2+4+21) also did not exhibit signal above the RNP control reaction (Figure 3C).

To determine if adding crRNA 21 would improve the limit of detection of our assay, we tested a combination reaction with crRNAs 2+4+21 on precisely titered SARS-CoV-2 genomic RNA obtained from the Biodefense and Emerging Infections Research Resources Repository (BEI Resources). In serial dilution experiments over 2 hours using 20 replicate reactions, the triple combination detected as few as 31 copies/μL (Figure 3D, left), based on the viral copy number independently determined by BEI with digital droplet (dd) PCR. By comparing the slope of an individual reaction with that of the RNP control, we determined that, for all dilutions, 20/20 individual tests (100%) would be correctly identified as “positive” with the 95% confidence level (Figure 3D, right).

Finally, we obtained five de-identified RNA samples extracted from nasal swabs taken from SARS-CoV-2+ individuals. Clinical RT-qPCR measurements resulted in Ct values of 14.37–22.13 for the patient samples, correlating to copy numbers 2.08 x 10^7^–1.27 x 10^5^ copies/μL. We added 0.3 μL of RNA from Patient Swabs 1-4 and 0.26 μL of RNA from Patient Swab 5 to each 20 μL Cas13a reaction (in triplicate). Using the direct detection assay on a plate reader, we correctly identified all five positive samples (ranging from 3.2 x 10^5^–1.65 x 10^3^ copies/μL in the Cas13a reaction), which showed slopes significantly above that of the RNP control (Figure 3E). The slopes correlated significantly with their input copy number (Pearson r coefficient = 0.9966, two-tailed p value = 0.0002), reinforcing the quantitative nature of the assay.

### Harnessing the Mobile Phone Camera as a Portable Plate Reader

To demonstrate that SARS-CoV-2 screening with Cas13a can be used outside of laboratory settings, we designed a mobile phone-based fluorescence microscope that measures the fluorescent signal generated by the Cas13a direct-detection assay (Figure 4A). Contrary to our expectation that a commercial laboratory plate reader would perform better than a mobile phone-based detection system, we found that our device was approximately an order of magnitude more sensitive than the plate reader due to reduced measurement noise and the ability to collect more time points, which decreased the uncertainty in slope estimations (Supplementary Figure 4).

**Figure 4:**
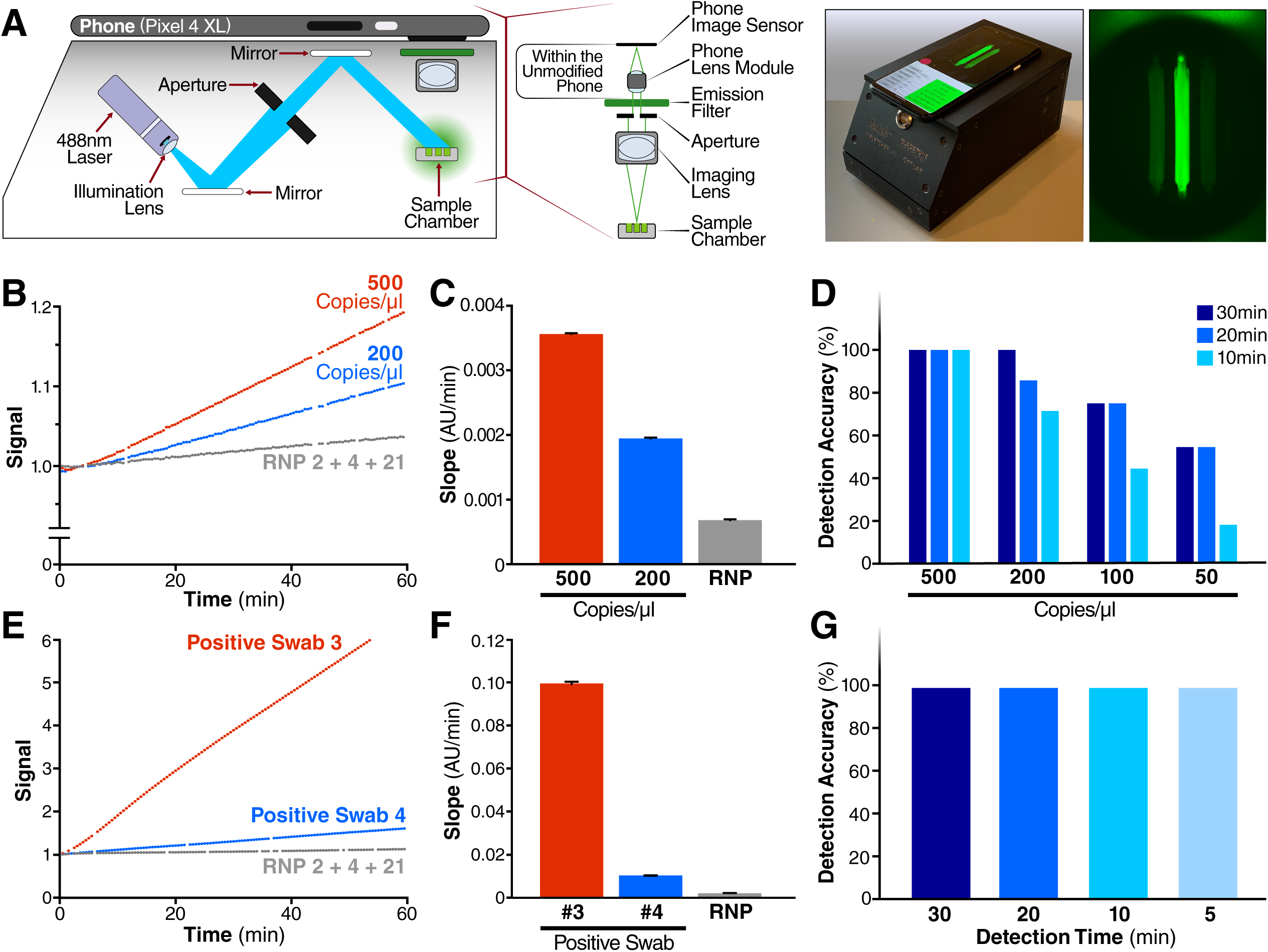
Harnessing the Mobile Phone Camera as a Portable Plate Reader. (A) Schematic of mobile phone-based microscope for fluorescence detection showing illumination and image collection components (left). Picture of assembled device used for data collection and sample image taken by the mobile phone camera after running a Cas13a assay (right). (B) Results from the Cas13a assay run on the mobile device with two different dilutions of full-length SARS-CoV-2 viral RNA isolated from infected Vero CCL81 cells (500 and 200 copies/µL) and RNP alone, using three combined crRNAs (crRNA 2, crRNA 4 and crRNA 21). Y-axis is the fluorescent signal of each sample normalized by the first time point. (C) Slope of the curve over 30 minutes from Figure 4B was calculated by simple linear regression and is shown as slope ± 95% confidence interval. (D) Detection accuracy of the Cas13a assay is characterized in the mobile device using SARS-CoV-2 full-length viral RNA. For each target dilutions, the slope at three different times - 10, 20 and 30 minutes - were compared to the slope of the no target RNA RNP alone controls, and the detection accuracy was determined at the 95% confidence level. The number of replicates for each concentration is 8 (500 copies/µL), 7 (200 copies/µL), 8 (100 copies/µL), and 11 (50 copies/µL). (E) Results from a Cas13a assay run on mobile device with two different nasal swab samples from human patients, confirmed as positive for SARS-CoV-2 using RT-qPCR, and the RNP alone control, all using the crRNA combination of crRNA 2, crRNA 4 and crRNA 21. (F) Slope of the curve over 30 minutes from Figure 4E was calculated by simple linear regression and is shown as slope ± 95% confidence interval. (G) Detection accuracy of Cas13a assay for n=5 nasal swab samples from human patients, confirmed as positive by RT-qPCR. Detection accuracy was evaluated at four different time points: 5, 10, 20 and 30 minutes.

We tested the performance of the device for detecting SARS-CoV-2 RNA using the triple-crRNA Cas13a assay and a dilution series with full viral RNA isolated from supernatants of infected Vero CCL-81 cells (Figures 4B-D). As with the plate reader, fluorescence generated in each reaction chamber was collected over time, with measurements every 30 seconds, showing a steady increase in fluorescence for full-length virus concentrations of 500–200 copies/μL, compared to RNP controls (Figure 4B). As before, the slopes of each curve can be calculated, along with the 95% confidence interval indicated by the error bars (Figure 4C). To determine the limit of detection of the direct detection assay on the device, 7-11 replicates each of dilution of virus corresponding to 500, 200, 100, and 50 copies/μL were measured, as determined by RT-qPCR. Slopes were calculated based on data for the first 30, 20, and 10 min of the assay, and each slope was then compared to the RNP control slope calculated over the same time. For each dilution and assay time, the ability of the assay to detect the target RNA relative to the RNP control was quantified as percent accuracy, with eight positive tests out of eight replicates for 500 copies/μL for all assay times corresponding to 100% accuracy (Figure 4D). The results over all dilutions indicate that the limit of detection is approximately 200 copies/μL in under 30 minutes, with accuracy dropping to 50% at 50 copies/μL.

Next, we analyzed the same patient samples as in Figure 3e to compare detection on the plate reader to that on the mobile phone-based device. We imaged each reaction for 60 minutes, along with the RNP control (Figure 4E), and the slope for a patient with Ct = 17.65 (Positive Swab 3) is significantly greater than the slope for a patient with Ct = 20.37 (Positive Swab 4) (Figure 4F), as expected. To assess the detection accuracy, we performed a linear fit using data from the first 5, 10, 15 and 20 minutes of the run and compared the slope of each sample to the RNP control. We determined all five samples to be positive within the first 5 minutes of the assay, indicating that the device can provide a very fast turnaround time of results for patients with clinically relevant viral loads (Figure 4G). This result highlights the inherent tradeoff between sensitivity and time in the Cas13a direct detection assay. High viral loads can be detected very rapidly because their high signals can be quickly determined to be above the control, and low viral loads can be detected at longer times once their signal can be distinguished above the control. In this way, our assay demonstrates a time-dependent sensitivity (Supplementary Figure 5) that can potentially be tuned to address both screening applications and more sensitive diagnostic applications.

## DISCUSSION

Here we show that direct detection of SARS-CoV-2 RNA with CRISPR-Cas13a and a mobile phone offers a promising option for rapid, point-of-care testing. A key advance in this work is demonstrating that combinations of crRNAs can increase the sensitivity of Cas13a direct-detection by activating more Cas13a per target RNA. We show that combinations of two or three crRNAs can be used to detect viral target RNA in the attomolar range or as few as ∼30 copies/μL. The use of multiple crRNAs that target different parts of the genome also safeguards against a potential loss of detection due to naturally occurring viral mutations.

A second key advance is the ability to directly translate the fluorescent signal into viral loads. Other CRISPR Dx assays, such as CRISPR-COVID, achieve high sensitivity via isothermal amplification but provide only qualitative information rather than information on viral copy number. Fluorescent signal from 7500 copies/μL to 7.5 copies/μL are remarkably similar, despite three orders of magnitude difference in copy number (Hou et al., 2020). By avoiding amplification and employing direct-detection, we show that the reaction rate directly correlates with viral copy number and may be used for quantification. When coupled with frequent testing, quantitative data are potentially beneficial: the course of a patient’s infection can be monitored, and one can determine if the infection is increasing or decreasing. In symptomatic cases, viral loads follow the course of the infection (Wolfel et al., 2020). Notably, samples with viral loads below 10^6^ copies/mL or 1000 copies/μL did not yield viral isolates in one study in Germany (Wolfel et al., 2020). Less is known about infectivity in asymptomatic cases, but SARS-CoV-2 transmission from asymptomatic patients has been documented (Bai et al., 2020), and asymptomatic patients may have viral RNA loads similar to those of symptomatic patients (Lee et al., 2020). Monitoring viral loads quantitatively will allow estimation of infection stage and will help predict infectivity, recovery and return from quarantine in real time.

A third key advance in our work is that a mobile phone-based device can accurately read the direct-detection assay, enabling ∼100 copies/μL sensitivity in 30 minutes and accurate diagnosis of a set of patient samples in 5 minutes. The device avoids the need for a bulky laboratory-based plate reader, makes the assay portable, and opens up the possibility for future point-of-care or at-home use. The choice of a mobile phone as the basis for our detection device was motivated by the high sensitivity of current mobile phone cameras, the simplicity of integrating a mobile phone for detection, their robustness and cost-effectiveness, and the fact that they are widely available today, meaning that they do not compete with other devices for limited production resources in the midst of a pandemic. For similar reasons, previous diagnostic efforts utilized mobile phones to detect fluorescent signals generated by molecular diagnostic, such as loop-mediated isothermal amplification (Chen et al., 2017; Ganguli et al., 2017; Kong et al., 2017; Priye et al., 2017; Sun et al., 2020), PCR (Angus et al., 2015; Gou et al., 2018; Jiang et al., 2014), next-generation DNA sequencing (Kuhnemund et al., 2017), and recombinase polymerase amplification reaction (Chan et al., 2018). Combined with efficient contact tracing and HIPAA-compliant upload into cloud-based systems, a mobile phone-based SARS-CoV-2 diagnostic could play an important role in control of the current and future pandemics.

The direct-detection assay reported here, if scaled up, could fulfill the need for a test that provides rapid results and is available to be administered frequently (Larremore et al., 2020). Other tests in this category include Abbott Lab’s ID NOW, a PCR-based test, and several antigen tests, such as Quidel’s Sofia 2 SARS Antigen FIA and Abbot Lab’s BinaxNOW Antigen Test. In the case of influenza, antigen tests span a wide range of sensitivities (e.g., 51–67.5%) (Babin et al., 2011; Chartrand et al., 2012; Chu et al., 2012). Due to the low-to-moderate sensitivity of these tests, the CDC still recommends re-testing samples that are negative with a more sensitive test, such as RT-qPCR (Green and StGeorge, 2018). Notably, none of the current rapid testing options provides quantitative results, which could help assess vial dynamics and evaluate an individual’s level of infection and progression of disease.

While we demonstrate rapid detection with reasonable sensitivity using crRNAs based on existing PCR primers, we anticipate further improvement by systematically searching for the best crRNA combinations across the entire viral RNA genome. As more information becomes available about viral variants (Osorio and Correia-Neves, 2020; Vanaerschot et al., 2020), crRNA design can be adapted to avoid false negatives or to specifically differentiate viral variants in the assay. Further improvements are also anticipated in the reporter, the choice of Cas13 orthologs and homologs, and in device and camera sensitivity. These advancements can improve the rate of the reaction, allowing for improvements in detection accuracy and limit of detection in shorter periods of time.

A recent national survey of over 19,000 respondents showed that the average wait time for nasal swab-based qPCR test results was 4.1 days, with 31% of tests taking more than 4 days and 10% of tests taking 10 days or more (Lazer et al., 2020). The national backlog in processing these laboratory-based tests clearly illustrates the need for rapid, point-of-care tests that can reliably detect SARS-CoV-2 RNA. As the long-term immunity induced by natural infection or vaccination may decay in as little as 2–4 months (Ibarrondo et al., 2020; Long et al., 2020), the need for rapid and frequent testing for SARS-CoV-2 will likely remain. In the future, direct detection by Cas13a as outlined here could be quickly modified to target the next respiratory pathogen that emerges, hopefully in time to help curb global spread.

## Data Availability

All data referred to in the manuscript is presented as such in the results, methods, or supplement.

## ACKNOWLEDGEMENTS

We thank all members of the Ott, Fletcher, and Doudna laboratories for sharing reagents, expertise and feedback throughout the preparation of this manuscript. We thank Google, in particular Michael Frumkin, Michael Brenner, Ellen Klein, Alex Schiffhauer, Daniel Harbuck, Francois Bleibel, Paul Rohde, Jon Barron, and Sam Hasinoff, for their assistance with this work and for donation of Pixel 4 phones. We thank Lauren Weiser and Veronica Fonseca for administrative support, Gary Howard for editorial support, and John Carroll for graphical support. We are very grateful to Kevin Mullane, Stephen B. Freedman, and Robert Wicks at the Gladstone Institutes, and JF Van Kerckhove, Savi Baveja, and Daniel LeClerc at Bain & Company for their guidance, support, and advice. We thank Paula Ladd and Cristina Tato for helpful discussions and advice. We thank the Gladstone Assay Development and Drug Discovery Core, specifically Michael Jobling, for assistance with high-throughput assays.

We gratefully acknowledge support from the NIH (grant 5R61AI140465-03 to J.A.D., D.A.F., and M.O.). This project is supported by the NIH Rapid Acceleration of Diagnostics (RADx) program and has been funded in whole or in part with Federal funds from National Heart, Lung and Blood Institute, National Institute of Biomedical Imaging and Bioengineering, National Institutes of Health, Department of Health and Human Services, under Grant No. 3U54HL143541-02S1. P.F. was supported by the UCSF Medical Scientist Training Program (T32GM007618) and the NIH/NIAID (F30AI143401). M.D.L.D was supported by the UC MEXUS-CONACYT Doctoral Fellowship. G.J.K. is supported by an NHMRC Investigator Grant (EL1, APP1175568) and previously an American Australian Association Fellowship. J.A.D. is an HHMI Investigator. We are grateful for philanthropic support from the James B. Pendleton Charitable Trust, Diksha and Divesh Makan, Kimberly and Stephen Richardson, Pamela and Edward Taft, and the Skoll Foundation. This work was made possible by a generous gift from an anonymous private donor in support of the ANCeR diagnostics consortium.

Purified LbuCas13a was a kind gift from Shanghai ChemPartner. The following reagents were deposited by the Centers for Disease Control and Prevention and obtained through BEI Resources, NIAID, NIH: Genomic RNA from SARS-Related Coronavirus 2, Isolate USA-WA1/2020, NR-52285; SARS-Related Coronavirus 2, Isolate USA-WA1/2020, NR-52281; and Human Coronavirus, NL63, NR-470.

## AUTHOR CONTRIBUTIONS

P.F., S.S., M.D.L.D, D.A.F., and M.O. conceived and designed the study. P.F., S.S., M.D.L.D., C.N.G., S.I.S., D.B., C.T., and J.S. performed experiments and data analysis. C.Z. and K.S.P. performed bioinformatics analysis. G.R.K. and J.M.O helped supervise and provide feedback. S.S., M.D.L.D., M.V.D., N.A.S., A.B, M.A., and A.H. developed the mobile phone-based device and reaction chambers. A.S.P provided HCoV-NL63 and HCoV-OC43 viral supernatants. C.L., E.D.C., M.P., A.K., and J.D.L provided patient samples and reagents. G.J.K. and J.A.D provided reagents, and critical expertise and feedback. D.A.F. and M.O. supervised the study design and data collection. J.A.D., D.A.F., and M.O. secured funding. P.F., S.S., M.D.L.D, D.A.F., and M.O. wrote the manuscript, with input from G.J.K. and J.A.D.

## METHODS

### Cas13a protein expression and purification

Expression vectors deposited with Addgene (Plasmid #83482) were used for expression of LbuCas13a. The codon-optimized Cas13a genomic sequences are *N-*terminally tagged with a His_6_-MBP-TEV cleavage site sequence, with expression driven by a T7 promoter. Purification of was based off of a previously published protocol with some modifications (East-Seletsky et al., 2017; East-Seletsky et al., 2016). Briefly, expression vectors were transformed into Rosetta2 DE3 or BL21 *E. coli* cells grown in Terrific broth at 37°C, induced at mid-log phase (OD600 ∼0.6) with 0.5 mM IPTG, and then transferred to 16°C for overnight expression. Cell pellets were resuspended in lysis buffer (50 mM Tris-Cl pH 7.0, 500 mM NaCl, 5% glycerol, 1 mM TCEP, 0.5 mM PMSF, and EDTA-free protease inhibitor (Roche)), lysed by sonication, and clarified by centrifugation at 35,000xg. Soluble His6-MBP-TEV-Cas13a was isolated over metal ion affinity chromatography, and in order to cleave off the His6-MBP tag, the protein-containing eluate was incubated with TEV protease at 4°C overnight while dialyzing into ion exchange buffer (50 mM Tris-Cl pH 7.0, 250 mM KCl, 5% glycerol, 1 mM TCEP). Cleaved protein was loaded onto a HiTrap SP column (GE Healthcare) and eluted over a linear KCl (0.25-1.0M) gradient. Cas13a containing fractions were pooled, concentrated, and further purified via size-exclusion chromatography on a S200 column (GE Healthcare) in gel filtration buffer (20 mM HEPES-K pH 7.0, 200 mM KCl, 10% glycerol, 1 mM TCEP) and were subsequently flash frozen for storage at -80°C.

### *In vitro* RNA transcription

SARS-CoV-2 N gene was transcribed off a single-stranded DNA oligonucleotide template (IDT). HCoV-MERS N gene was transcribed off of a MERS-CoV Control plasmid (IDT, Cat # 10006624) by first adding a T7 promoter via PCR using Q5 High-Fidelity DNA Polymerase (NEB) (see table for primers). A single PCR product was confirmed via gel electrophoresis. *In vitro* transcription was performed using HiScribe T7 Quick High Yield RNA Synthesis Kit (NEB) following manufacturer’s recommendations. Template DNA was removed by addition of DNase I (NEB), and IVT RNA was subsequently purified using RNA STAT-60 (AMSBIO) and the Direct-Zol RNA MiniPrep Kit (Zymo Research). RNA concentration was quantified by Nanodrop and copy number was calculated using transcript length and concentration.

### SARS-CoV-2 virus culture

Isolate USA-WA1/2020 of SARS-CoV-2 was used for full-length viral RNA. All live virus experiments were performed in a Biosafety Level 3 laboratory. SARS-CoV-2 stocks were propagated in Vero CCL-81 cells. Viral supernatant was collected by centrifugation for RNA extraction (see below).

### HCoV-NL63 virus culture

Isolate Amsterdam I of HCoV-NL63 (NR-470, BEI Resources) was propagated in Huh7.5.1-ACE2 cells. Supernatant was harvested 5 days post infection, filtered and stored at -80C.

### HCoV-OC43 virus culture

HCoV-OC43 (VR-1558, ATCC) was propagated in Vero E6 cells. Supernatant was harvested 6 days post infection, filtered and stored at -80C.

### Influenza virus

H1N1 Influenza virus A (California/04/2009) and Influenza virus B (Brisbane/60/2008) in chicken allantoic fluid was purchased from Virapur and used directly for RNA extraction (see below).

### RNA extraction

RNA was extracted from SARS-CoV-2, HCoV-NL63, HCoV-OC43, Influenza A, and Influenza B viral supernatant via RNA STAT-60 (AMSBIO) and the Direct-Zol RNA MiniPrep Kit (Zymo Research). RNA was extracted from human airway organoid cells using the RNeasy Mini Kit (Qiagen).

### Quantitative polymerase chain reaction

RNA from SARS-CoV-2 viral supernatant was quantified via qPCR. Briefly, RNA was reverse transcribed to cDNA via AMV Reverse Transcriptase (Promega) using oligo(dt)18 and random hexamers (Thermo Scientific). cDNA was added to the qPCR reaction using PrimeTime Gene Expression Master Mix (IDT). N and E gene standards were used to generate a standard curve for copy number quantification. N gene standard was generated by PCR from the 2019-nCoV_N_Positive Control Plasmid (IDT, Cat #10006625). E gene standard was generated by PCR using extracted full-length SARS-CoV-2 RNA as template. A single product was confirmed by gel electrophoresis and DNA was quantified by Nanodrop. cDNA was analyzed using the 7500 Fast Real-Time PCR system (Applied Biosystems). See table for primers.

### Fluorescent Cas13a nuclease assays

LbuCas13a-crRNA RNP complexes were individually preassembled by incubating 1.33 μM of LbuCas13a with 1.33 μM of crRNA for 15 minutes at room temperature. In Figures 1C and 2B, 677 nM of crRNA was used. These complexes were then diluted to 100 nM LbuCas13a and 100 nM (or 50 nM for Figures 1C and 2B) crRNA in cleavage buffer (20 mM HEPES-Na pH 6.8, 50 mM KCl, 5 mM MgCl2, and 5% glycerol) in the presence of 400 nM of reporter RNA (5’-FAM-rUrUrUrUrU-IowaBlack FQ-3’), 1 U/μL Murine RNase Inhibitor (NEB, Cat #M0314), and varying amounts of target RNA. In Figures 1C and 2B, 167 nM of RNase Alert substrate (IDT) was used as the reporter RNA, and in Figure 2D, 400 nM of RNase Alert substrate was used. In Figure 3D and 3E, the complexes were also diluted in 0.1% Tween-20 (Sigma) and the cleavage buffer was pH 7.1. These reactions were incubated in a fluorescence plate reader for up to 120 minutes at 37°C with fluorescence measurements taken every 5 minutes (or every 2.5 minutes in Figure 3E) (λ_ex_:485 nM; λ_em_:535 nM). Background-corrected fluorescence values were obtained by subtracting fluorescence values obtained from reactions carried out containing only reporter and buffer. For assays containing more than one crRNA simultaneously, the LbuCas13a-crRNA RNP complexes were separately assembled by incubating for 15 minutes at room temperature, then combined in the reaction at half (in 2 RNP combinations) or one-third (in 3 RNP combinations) the volume to keep the total or combined concentration of RNP constant. Representative graphs of experiments are shown. Most experiments were replicated at least twice, with the exception of Figure 3E (only Patient Swabs 1-3 and 5 were repeated twice) due to limited sample material.

### Patient Samples

De-identified RNA samples from nasopharyngeal swabs from patients testing positive and negative for SARS-CoV-2 were obtained from the Chan Zuckerberg Biohub. Positive samples were quantified previously using CDC N1 and N2 SARS-CoV-2 primers.

### Mobile phone fluorescent microscope

We built a mobile phone fluorescent microscope using a 488 nm diode laser (Sharp 55mW, DTR’s Laser Shop), a green fluorescence interference filter (Chroma Technology AT 535/40), and a Pixel 4 XL phone camera (12.2 mega-pixel, pixel size 1.4μm, aperture f/1.7, Google). The laser beam was expanded using a glass collimation lens (10° divergence half-angle) (DTR-G-8, DTR’s Laser Shop), directed towards the sample plane using two ND4 filters used as mirrors (ND40B, Thorlabs), and reduced by an elliptical aperture to fill the circular image field-of-view with a uniform field intensity. The sample was illuminated with an oblique epi configuration and the illumination power was 18 mW at the sample plane. The imaging optics consist of an f=20mm compact triplet lens (TRH127-020-A, Thorlabs) followed by the interference filter for selection of the fluorescence reporter emission wavelength and the Pixel 4 XL camera lens. Total magnification from object to image plane is 1/3 and the numerical aperture is 0.06. All optical and illumination components were enclosed in a custom-made dark box, into which a sample chip is loaded for imaging. Automated time-lapse imaging was implemented by a custom Android application and a Bluetooth receiver (Bluefruit Feather M0, Adafruit), which triggered the laser at the time of image acquisition. The Cas13a reaction was performed by placing the device in a 37°C incubator for temperature control and the reaction curve was obtained by analyzing the image time series offline using a custom MATLAB (Mathworks) code.

### Sample chip fabrication

Sample chips containing three fluid channels were made by casting polydimethylsiloxane (PDMS, Ellsworth adhesives) on an acrylic mold. The acrylic mold was assembled by adhering three laser cut acrylic lanes on a flat acrylic base. The width, height, and length of each acrylic lane were 2 mm, 2 mm, and 10 mm, respectively, resulting in a fluid channel volume of 40 µL. Inlet and outlet ports were created on both ends of the channels after curing and demolding the PDMS using a biopsy punch. The PDMS chips were subsequently adhered to a siliconized cover glass (Hampton research) to close the fluid channels. To avoid generation of bubbles in the chip during the measurement, both the Cas13a reaction mix and the sample chip were degassed in a house vacuum for 15 minutes before loading the samples and starting the measurement.

### Mobile phone image acquisition and analysis

During typical device operation, an RGB image was acquired every 30 seconds for the duration of 1 hour and the images were analyzed offline using a custom MATLAB code. First, the RGB image was demosaiced to a greyscale image. Second, the saturated pixels or pixels exhibiting two very different green submosaic values were excluded. Third, a rectangular image region-of-interest (ROI) was drawn within an area of each fluid channel and the reporter signal in each ROI was determined by averaging the pixel values.

### Quantification and statistical analysis

Data in Figures 1-3 were processed and visualized using GraphPad Prism.

**Table 1:**
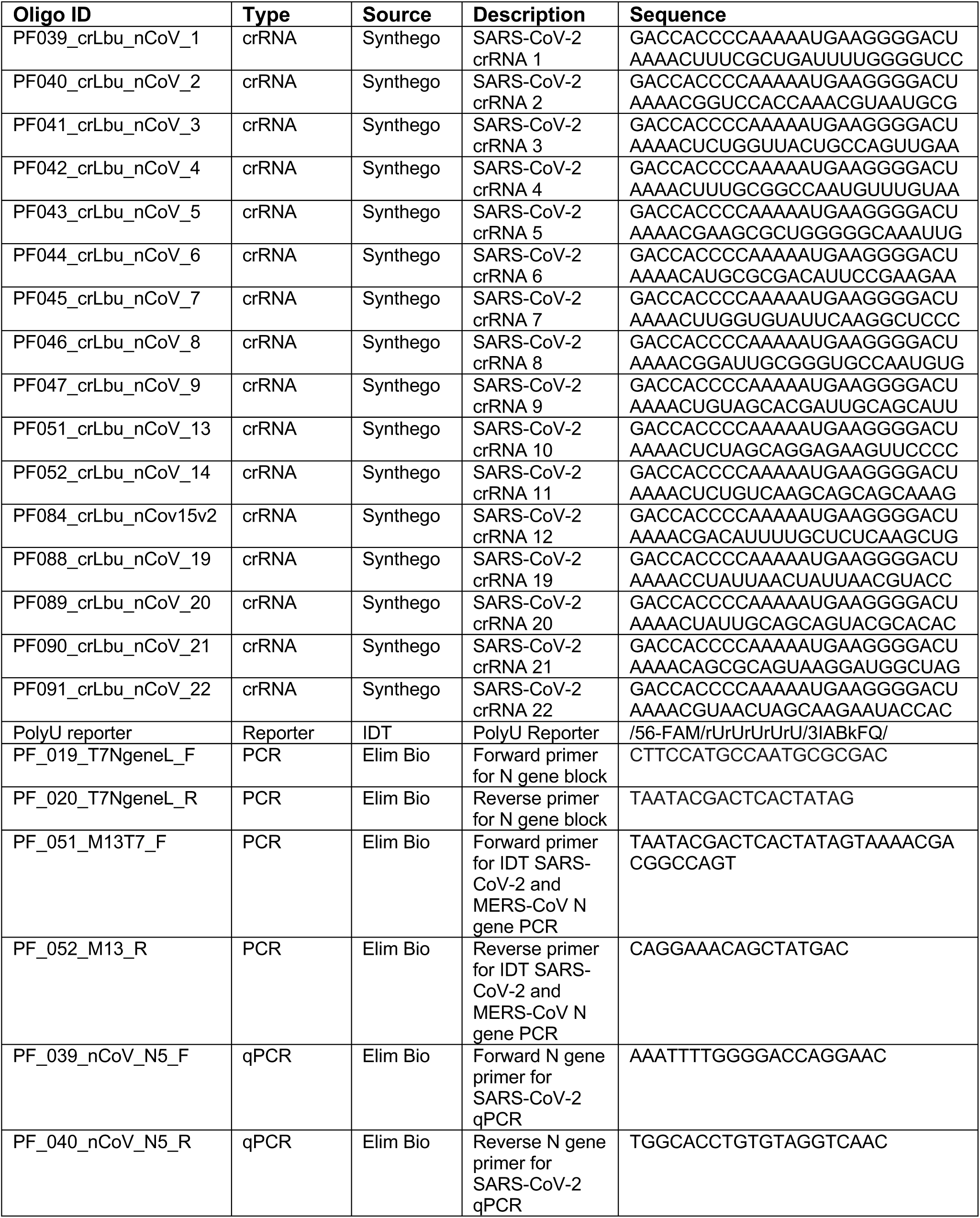
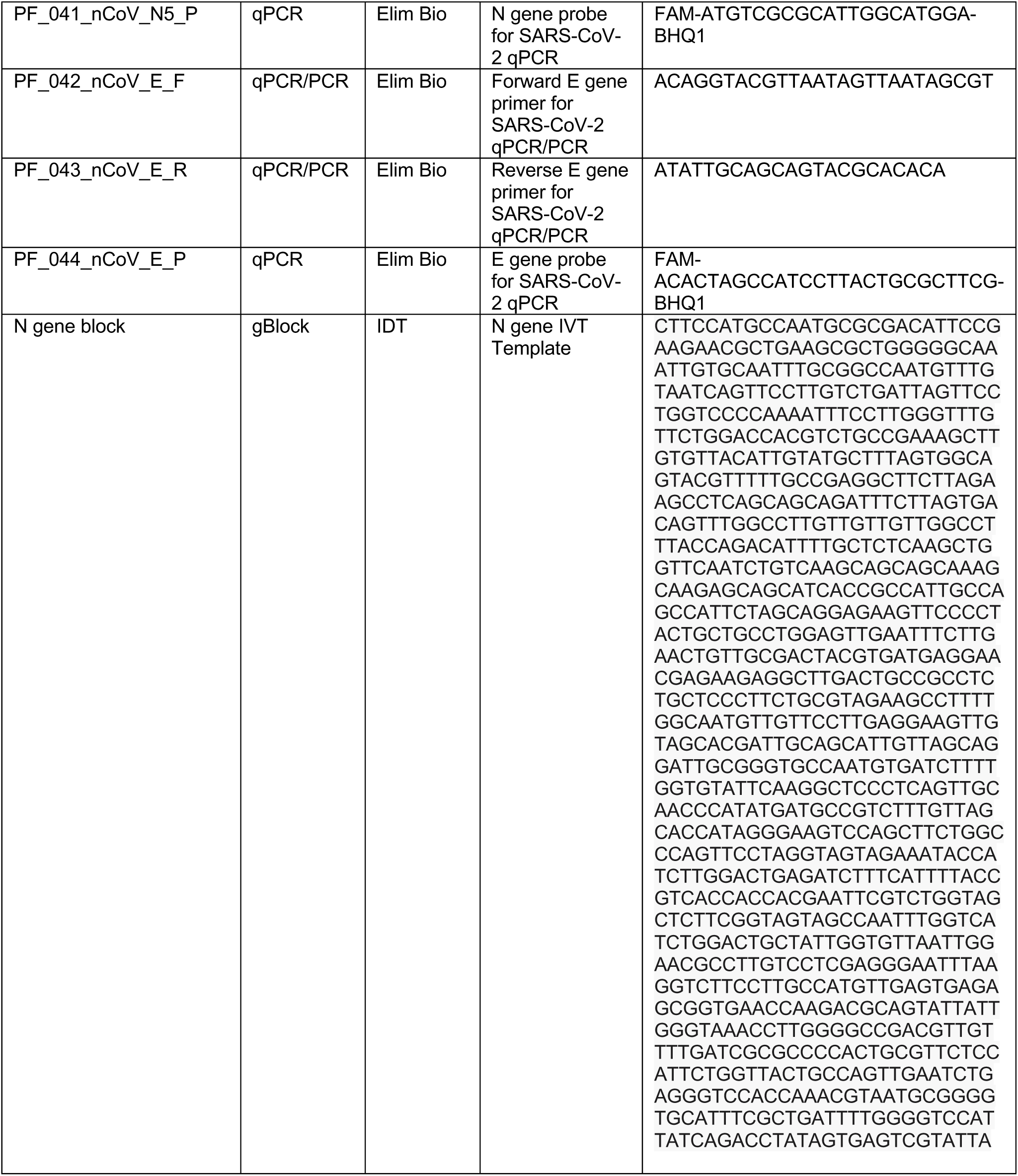
List of custom oligonucleotides used in this study

